# Operationalising social transition in trans and gender diverse youth: a scoping review of definitions and measures

**DOI:** 10.1101/2025.10.27.25338426

**Authors:** Will Conabere, Francesca Lami, Hannah Andrews, Mary Skarlatos, Carmen Pace, Pip Buckingham, Sam Hay, Tahlia De Ieso, Ken C. Pang, Michelle Tollit

## Abstract

**Background:** Social transition can be an important and accessible aspect of gender affirmation, allowing trans and gender diverse young people to authentically express their identities, including through adoption of chosen name, pronouns, and changes to appearance. These changes may take place in a single context, such as at home, or across multiple settings, including school, with friends, and online. Currently, it is unclear how this process is being captured in the literature, including whether it has been measured comprehensively and consistently.

**Methods:** This scoping review explored how social transition has been defined and operationalised in studies involving trans young people (under 18), detailing practices (i.e. specific changes) of social transition and contexts where these changes occurred. Systematic searches were conducted in Embase, MEDLINE, and PsycInfo databases. Studies were eligible if they included a definition and/or measure of social transition.

**Findings:** Among 40 included studies, definitions (n=37 studies) and measures (n=29 studies) varied widely. Some studies did not distinguish between specific practices of social transition, such as changes to name, pronouns, or appearance, when defining (5/37) and measuring (8/29) the construct. Similarly, many studies omitted reference to the contexts in which these practices occurred (18/37 definitions; 11/29 measures), assuming practices took place uniformly across all contexts. Many also relied on a binary measure of social transition (15/29 measures), failing to capture the diverse ways in which individuals enact their social transition. No measurement tools were used consistently.

**Interpretation:** Current definitions and measures of social transition in research with trans and gender diverse young people are inconsistent and incomplete. As a result, they fail to capture the complex nature of social transition, limiting comparability between studies and hindering understanding of its complexities and impacts. Comprehensive, standardised measures to capture this concept are urgently required.

**Funding:** There was no funding source for this study.

**Research In Context:** *Evidence before this study:* This scoping review examines how social transition, i.e., the process by which transgender and gender diverse people outwardly express their gender identity, has been operationalised in the literature regarding young people. While social transition is often described/reported to be complex and fluid, most studies appear to overlook this complexity. To date, no review has systematically and quantitatively assessed: (i) how social transition has been defined in empirical studies, or (ii) the breadth, depth, and consistency of the measures used to capture social transition. To address these evidence gaps, we searched Embase, MEDLINE and PsycInfo databases until September 14^th^, 2024, using terms relating to transgender identity (e.g. “transgender” or “gender diverse”), children and adolescents (e.g. “child” or “adolescent” or “adolescence”) and social transition (e.g. “social transition” or “pronoun” or “name”). From the included studies, we extracted information on how social transition was defined and measured. We focused on the extent to which studies recognised the specific practices (i.e. changes) involved, and the contexts in which these changes may occur (which may differ between individuals based on underlying levels of safety and support).

*Added value of this study:* This scoping review identified significant variability and oversimplification in how social transition has been defined and measured in studies with trans and gender diverse young people. Key aspects of social transition, such as the contexts in which changes occur, desire to make social transition changes, and some specific practices, were frequently underrepresented. Measures were also often based on narrow or binary constructs, applied inconsistently across studies, limiting their accuracy and comparability.

*Implications of all the available evidence:* By highlighting gaps in current measures of social transition, this study provides a foundation for future research to adopt more accurate and meaningful approaches for understanding and measuring social transition in line with the real-world experiences of transgender and gender diverse young people. Ensuring consistent measurement is essential to strengthen the evidence base and improve comparability and to support future research to accurately guide policy and clinical care for trans and gender diverse young people worldwide.

## Introduction

Transgender and gender diverse children and adolescents are those whose gender identity differs from the sex assigned to them at birth.^1^ Recent estimates suggest that approximately 1.2-2.7% of young people identify as transgender, within a broader 2.5-8.4% who identify as gender diverse.^1^ Affirming one’s gender identity can involve a range of processes across several contexts, including medical and non-medical interventions.

The process of social transition encompasses a range of non-medical gender-related outward changes that allow trans individuals to express their gender more authentically.^2^ These changes can include adopting a new name, using different pronouns, altering one’s appearance (e.g. hairstyle and clothing), modifying communication styles (e.g. voice), and participating in traditionally gender-aligned activities.^3–5^ Social transition can unfold quickly or gradually, as individuals may introduce these changes progressively across different contexts or with various groups, such as at home, among friends, at school or online.^6^ For many young people, social transition may be the most accessible or preferred avenue of gender affirmation. Limited access to gender-affirming medical care, whether due to jurisdictional limitations, long waitlists, or a lack of parental support, can present barriers to medical and legal affirmation for trans children and adolescents.^1,7^ It is also important to acknowledge that some individuals may have no desire for medical or legal interventions, progressing in their gender journey through social changes alone.^1^ Additionally, for pre-pubescent trans children, social transition is frequently used as the sole avenue of affirmation.^1^

Despite many trans people undertaking social transition without clinical input, social transition has received increased attention and scrutiny from some health professionals in recent years, particularly for young people. For example, the Cass Review recommended that social transition in children and adolescents should only occur under clinical supervision.^8^ This controversial recommendation has been widely criticised, not only for medicalisation of this social process^9^ and misinterpreting findings,^9–11^ but also for treating social transition as a uniform experience with no regard for how the concept was operationalised across their cited research. Given the numerous practices that can comprise social transition, evaluating how it is being captured is essential for accurate interpretation. However, to date, no studies have systematically explored how social transition is being characterised in the literature regarding young people.

Given the increased attention to social transition among transgender and gender diverse young people and the lack of clarity on how this process is conceptualised in research, this scoping review aimed to explore definitions and measures of social transition being used with transgender and gender diverse participants under 18 years. In doing so, we seek to provide a summary of the breadth, comprehensiveness, and consistency of existing approaches used to define and operationalise social transition in research with children and adolescents.

## Methods

### Protocol and registration

The protocol for this review is available on the PROSPERO database (PROSPERO 2022 CRD42022333058), where it was initially conceptualised as a systematic review. During the review process, the study was reframed as a scoping review to better align with its aims of capturing definitions and measures of social transition.^12^ The methods are reported in accordance with the Preferred Reporting Items for Systematic reviews and Meta-Analyses extension for Scoping Reviews (PRISMA-ScR).^13^

### Eligibility criteria

Quantitative and qualitative studies were eligible for inclusion. Studies were included if they (1) defined or measured (using either a questionnaire or a single item) social transition with children and adolescents (aged <18 years), and (2) at least 75% of participants in the study were under the age of 18 years. To ensure inclusivity of diverse gender identities, study selection was not based on the gender identities present in the sample. No date range or geographical location criteria were applied during the search and all non-English papers were excluded.

### Search strategy

Controlled vocabulary (i.e. MeSH terms) and keywords that captured terms relating to *transgender*, *children and adolescents* and *social transition*, were combined using Boolean operators and searched in MEDLINE, EmBase, and PsycINFO databases, covering the literature from 1946 to 2024 (Supplementary material 1). These databases were chosen to broadly cover biomedical, clinical, and psychological sciences, where literature on our topic of interest is likely to exist. The search was run on three occasions, the first search was conducted in May 2022 and updated with a second and third search in September 2024 and August 2025, respectively.

### Study selection

Duplicate records were identified and removed either manually or using automated procedures in Endnote^14^. Publications garnered through the electronic database search were then independently screened for eligibility by two pairs of reviewers for each phase of the study (first search: HA, MS; subsequent searches: WC and MS or SH). Full texts of eligible studies after title and abstract screening were then retrieved and independently assessed for inclusion by two pairs of reviewers (first search: MS and FL; subsequent searches: WC and SH). Any disagreements were resolved through discussion and, if required, a third reviewer was consulted. A snowballing technique from included papers was also used (Figure 1).

**Figure 1:**
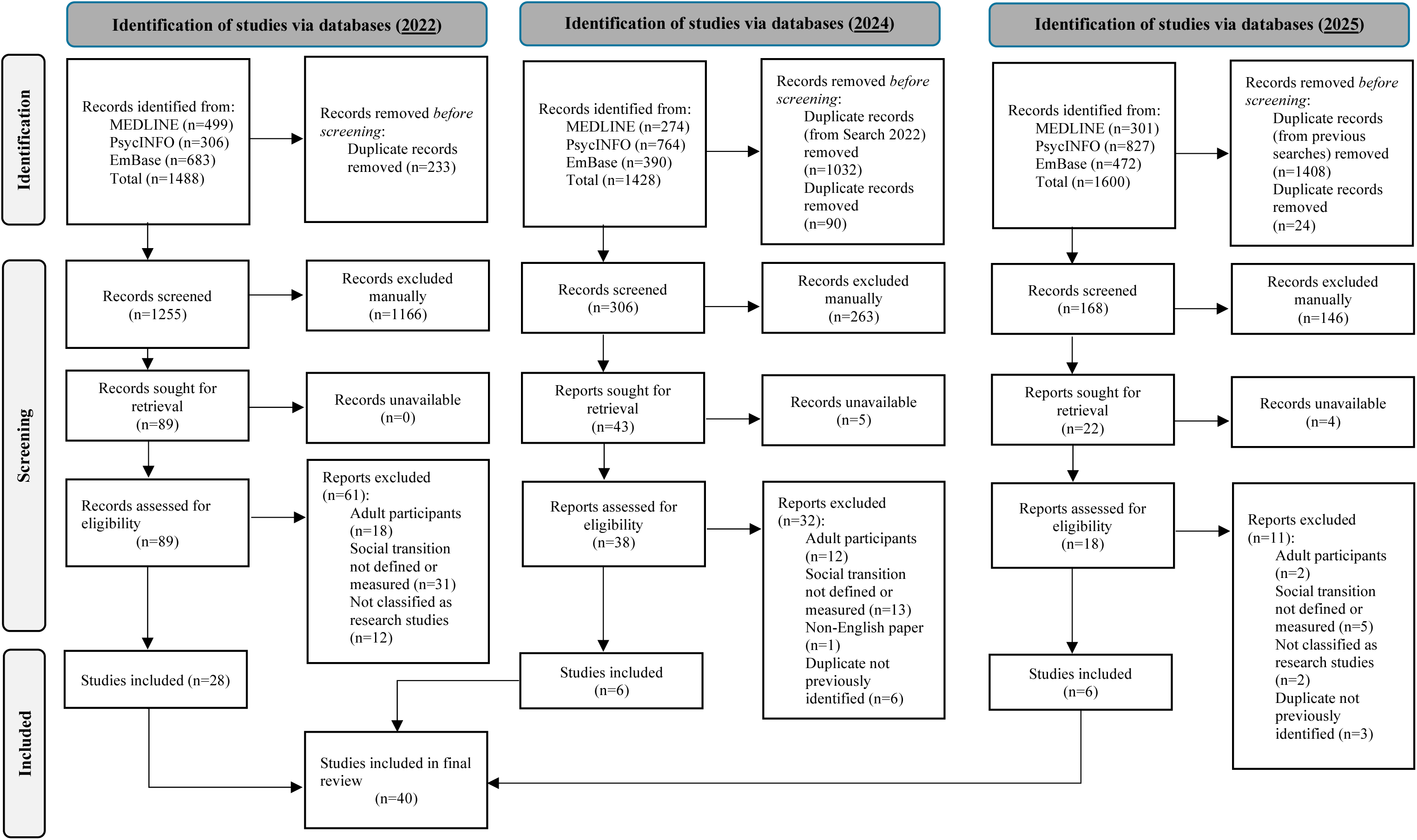
PRISMA Chart showing the study selection process.

### Data extraction

Data from included studies were extracted using standardised REDCap forms^15^ by two authors independently (FL, MS, WC, SH) to ensure accuracy. Any inconsistencies were resolved through discussion and, if required, a third reviewer was available for consultation. Data extracted included study characteristics (e.g. study design, age range(s) of participants, sample size, and study aims), as well as details related to the definition and/or measure of social transition. The concept of “definition” was identified as any explicit description of what social transition referred to in the study, as stated by the authors of the paper, and data extracted included the definition provided, practices of social transition referenced (e.g. name, pronouns) and contexts referenced (e.g. home, school). The concept of “measure” was referred to the operationalisation of social transition using specific items, checklists, or survey instruments that were used to capture transition-related practices. In terms of measures, data extracted included the number of participants to whom the measure was administered, the informant(s) (e.g. self-report, parent-report), how social transition was assessed (e.g. survey, interview), the nature of the scale (e.g. binary, dimensional, ordinal, other), the name of the measure (if applicable), and the practices of social transition (specific changes) and contexts assessed in the measure.

### Data synthesis

Definitions and measures of social transition were extracted, and the practices, contexts, number of practices, and number of contexts were summarised. Tabular summaries provided an overview of the breadth and consistency across studies, allowing for comparisons within studies (i.e. definition vs measure) and between studies.

To contextualise the included literature, studies were also categorised into six broad aim-based groups: (1) studying the association of social transition and other outcomes, (2) comparing differences in one or more variables between two or more groups (including a group of socially transitioned trans young people), (3) describing the process of social transition, (4) studying the effect of social transition in the association between two other variables, (5) studying the association between two variables in a group of socially transitioned young people, and (6) other aims not fitting the aforementioned categories. Studies that used social transition as an inclusion criterion were included in the measures analysis only if the criterion was explicitly operationalised in text. If the inclusion criterion was not defined, these studies were excluded from the measures analysis. Eligible inclusion criterion measures were categorised as “binary measures”, as the recorded data only indicated whether participants met or did not meet the criteria, without detailing which specific aspects of social transition were met/not met.

### Critical appraisal

In line with expectations for scoping reviews,^16^ assessment of risk of bias were not conducted. This aligns with the review’s focus on understanding how social transition is defined and measured, rather than evaluating data derived from the included studies.

### Positionality Statement

The authors acknowledge that their perspectives are shaped by their professional backgrounds in trans health and public health as well as by their positionalities as both cisgender and gender diverse clinicians and researchers. The diversity of our team informed our commitment to conducting this work with sensitivity to the lived experiences of trans and gender diverse young people.

## Results

### Study selection

The literature searches identified 1729 unique titles/abstracts for screening, leaving 145 full text papers to be assessed, after which a total of 40 studies were included in the final review (Figure 1). Data obtained from these studies (i.e. definitions and measures) are presented in Supplementary Table 1.

### Study characteristics

Table 1 presents key study characteristics, including the study design, basic participant demographics and research focus of the 40 included studies. The majority of these studies were quantitative (n=32, 80·0%), with 27 cross-sectional and five longitudinal. Data on social transition were provided by trans and gender diverse young people in 19 studies, their parent or caregiver in 11 studies, both (parent and child) in seven studies or obtained using other methods (e.g. medical records) (n=3).

**Table 1:** Demographic information for each included study.

| First author, year | Country | Study design | Total Participants (n) | Child Age Range (years) | Focus of investigation |
| --- | --- | --- | --- | --- | --- |
| Aramburu-Algeria, 2018<sup>17</sup> | United States | Qualitative | 14 | 6-17 | Exploratory qualitative study (not on social transition) |
| Bocek, 2024<sup>18</sup> | United States | Quantitative | 294 | 13-17 | Association between two non-social transition outcomes |
| Bull, 2018<sup>19</sup> | United States | Qualitative | 8 | 4-12 | Exploratory qualitative study (on social transition) |
| De Castro, 2024<sup>20</sup> | Spain | Quantitative | 140 | Up to 17 | Association between social transition and another outcome |
| Durwood, 2017<sup>21</sup> | North America* | Quantitative | 179 | 6-14 | Association between two non-social transition outcomes or social transition and another outcome (unclear) |
| Durwood, 2021<sup>22</sup> | North America* | Quantitative | 265 | 3-15 | Association between two non-social transition outcomes |
| Durwood, 2022<sup>23</sup> | North America* | Qualitative | 14 | 6-14 | Exploratory qualitative study (on social transition) |
| Durwood, 2024<sup>24</sup> | North America* | Quantitative | 51 | 3-12 | Association between social transition and another outcome |
| Engel, 2023<sup>25</sup> | Australia | Quantitative | 525 | 6-17 | Association between social transition and another outcome and Association between two non-social transition outcomes |
| Fast, 2018<sup>26</sup> | United States | Quantitative | 96 | 3-5 | Association between social transition and another outcome |
| Gonzalez-Mendiondo, 2024<sup>27</sup> | Spain | Qualitative | 22 | 4-18 | Exploratory qualitative study (on social transition) |
| Gulgoz, 2022<sup>28</sup> | North America* | Quantitative | 300 | 3-14·58 | Association between social transition and another outcome and Association between two non-social transition outcomes |
| Haywood, 2023<sup>6</sup> | United Kingdom | Quantitative | 81 | 9-17 | Association between social transition and another outcome |
| Horton, 2023<sup>29</sup> | United Kingdom | Qualitative | 30 | 6-16 | Exploratory qualitative study (not on social transition) |
| Kuper, 2019<sup>30</sup> | United States | Quantitative | 224 | 6-17 | Comparing participants by social transition status and association between social transition and another outcome |
| Kuvalanka, 2014<sup>31</sup> | United States | Qualitative | 5 | 8-11 | Exploratory qualitative study (not on social transition) |
| Kuvalanka, 2017<sup>32</sup> | United States | Quantitative | 45 | 6-12 | Descriptive quantitative study (not on social transition) |
| Legrange, 2024<sup>33</sup> | France | Quantitative | 239 | 3-18 | Association between social transition and another outcome |
| Lane, 2025<sup>34</sup> | United Kingdom | Quantitative | 1512 | 3-13 | Association between two non-social transition outcomes |
| Levitan, 2019<sup>35</sup> | Germany | Quantitative | 180 | 11-18 | Association between social transition and another outcome |
| Morandini, 2022<sup>36</sup> | United Kingdom | Quantitative | 782 | 4-17 | Descriptive quantitative study (not on social transition) |
| Morandini, 2023<sup>37</sup> | United Kingdom | Quantitative | 357 | 4-17 | Association between social transition and another outcome |
| O'Bryan, 2018<sup>38</sup> | United States | Quantitative | 139 | 8-21 | Association between two non-social transition outcomes |
| Olson, 2016<sup>39</sup> | North America* | Quantitative | 73 | 3-12 | Comparing participants by social transition status |
| Olson, 2018<sup>40</sup> | North America* | Quantitative | 149 | 6-8 | Association between social transition and another outcome |
| Olson, 2019<sup>41</sup> | North America* | Qualitative | 120 | 3-14 | Exploratory qualitative study (on social transition) |
| Olson, 2022<sup>42</sup> | North America* | Quantitative | 317 | 3-12 | Descriptive quantitative study on social transition over time |
| Rae, 2019<sup>43</sup> | North America* | Quantitative | 85 | 5-11 | Comparing participants by social transition status and association between social transition and another outcome |
| Renley, 2022<sup>44</sup> | United States | Quantitative | 4000 | 13-17 | Association between social transition and another outcome |
| Schimmel-Bristow, 2018<sup>45</sup> | United States | Qualitative | 18 | 14-22 | Exploratory qualitative study (on social transition) |
| Shim, 2020<sup>46</sup> | United States | Quantitative | 35 | 13-16.8 | Descriptive quantitative study (not on social transition) |
| Sievert, 2021<sup>47</sup> | Germany | Quantitative | 54 | 5-11 | Association between social transition and another outcome |
| Sood, 2021<sup>48</sup> | United States | Quantitative | 156 | 12-18 | Association between social transition and another outcome |
| Sorbara, 2020<sup>49</sup> | Canada | Quantitative | 300 | 10.5-17.9 | Association between two non-social transition outcomes |
| Steensma, 2013<sup>50</sup> | Netherlands | Quantitative | 127 | 6-12 | Descriptive quantitative study (not on social transition) |
| Thoma, 2023<sup>51</sup> | United States | Quantitative | 1943 | 14-18 | Association between social transition and another outcome |
| Tollit, 2024<sup>52</sup> | Australia | Quantitative | 522 | Age range not provided | Association between social transition and another outcome |
| van der Vaart, 2022<sup>53</sup> | Netherlands | Quantitative | 312 | 7-13 | Comparing participants by social transition status |
| Wittlin, 2025<sup>54</sup> | North America* | Quantitative | 183 | 3-12 | Comparing participants by social transition status |
| Wong, 2019<sup>55</sup> | North America* | Quantitative | 266 | 3-12.92 | Comparing participants by social transition status and association between social transition and another outcome |

\*North America = United States and Canada

Participant ages ranged from 3 to 22 years, with all studies reporting a mean participant age below 18 years. Fifteen studies included only children (<13 years), six included only adolescents (>12 years) and eighteen included both children and adolescents. Sample sizes ranged from 5 to 60 participants in qualitative studies, and 35 to 4,000 participants in quantitative studies. Among quantitative studies, the median sample size was 174 (IQR=208). The included studies had varied aims, with the most common focus (18/40 studies) being social transition in relation to another outcome. Seven other studies explored associations between outcomes unrelated to social transition. Among the eight qualitative studies, five investigated social transition as a primary focus.

### Definition of social transition

Definitions of social transition are presented in Supplementary Table 2 (n=37), while Tables 2, 3 and 4 summarise the nature of these definitions, including specific practices relating to social transition and contexts in which these practices have occurred. Of the 37 definitions, 30 were explicitly stated within the studies, while in seven cases, the definition of social transition was inferred from the description of the measure. In three instances, no definition could be inferred.

**Table 2:**
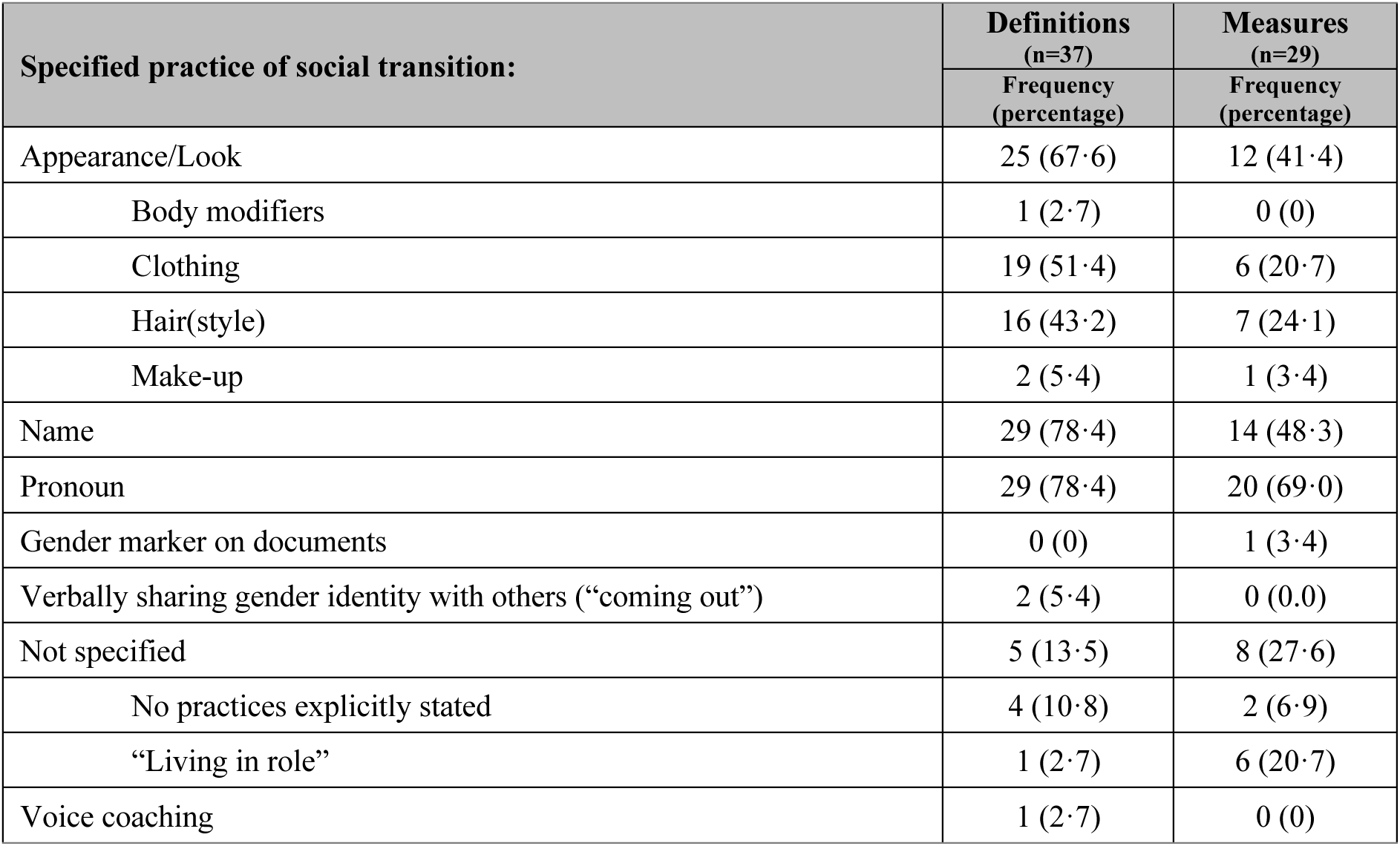
Frequency of social transition practices specified in definitions and measures.

| Specified practice of social transition: | Definitions<br>(n=37) | Measures<br>(n=29) |
| --- | --- | --- |
|  | Frequency<br>(percentage) | Frequency<br>(percentage) |
| Appearance/Look | 25 (67·6) | 12 (41·4) |
| Body modifiers | 1 (2·7) | 0 (0) |
| Clothing | 19 (51·4) | 6 (20·7) |
| Hair(style) | 16 (43·2) | 7 (24·1) |
| Make-up | 2 (5·4) | 1 (3·4) |
| Name | 29 (78·4) | 14 (48·3) |
| Pronoun | 29 (78·4) | 20 (69·0) |
| Gender marker on documents | 0 (0) | 1 (3·4) |
| Verbally sharing gender identity with others (“coming out”) | 2 (5·4) | 0 (0·0) |
| Not specified | 5 (13·5) | 8 (27·6) |
| No practices explicitly stated | 4 (10·8) | 2 (6·9) |
| “Living in role” | 1 (2·7) | 6 (20·7) |
| Voice coaching | 1 (2·7) | 0 (0) |

**Table 3:** Frequency of social transition contexts specified in definitions and measures.

| Specified context of social transition: | Definitions<br>(n=37) | Measures<br>(n=29) |
| --- | --- | --- |
|  | Frequency<br>(percentage) | Frequency<br>(percentage) |
| All aspects (contexts) of their life | 2 (5·4) | 3 (10·3) |
| Friends/peers |  |  |
| With friends | 3 (8·1) | 4 (13·8) |
| With peers | 1 (2·7) | 0 (0) |
| Family |  |  |
| Home (or with parents) | 13 (35·1) | 11 (37·9) |
| With extended family | 2 (5·4) | 1 (3·4) |
| Community |  |  |
| Bathrooms/Locker rooms | 1 (2·7) | 1 (3·4) |
| In public | 3 (8·1) | 1 (3·4) |
| With strangers | 1 (2·7) | 2 (6·9) |
| Not specified | 19 (51·4) | 12 (41·4) |
| Online | 1 (2·7) | 3 (10·3) |
| On official or legal documents | 0 (0) | 1 (3·4) |
| Private life | 1 (2·7) | 1 (3·4) |
| School/work |  |  |
| School | 12 (32·4) | 13 (44·8) |
|  | Frequency<br>(percentage) | Frequency<br>(percentage) |
| With teachers | 1 (2·7) | 0 (0) |
| At work | 2 (5·4) | 2 (6·9) |

**Table 4:** Number of specified practices and contexts in definitions and measures of social transition.

| Number of practices/contexts specified (per study): | Definitions<br>(n=37) | Measures<br>(n=29) |
| --- | --- | --- |
|  | Frequency<br>(percentage) | Frequency<br>(percentage) |
| **Practices** |  |  |
| Zero | 4 (10·8) | 2 (6·9) |
| One | 4 (10·8) | 12 (41·4) |
| Two | 3 (8·1) | 4 (13·8) |
| Three | 7 (18·9) | 7 (24·1) |
| Four | 14 (37·8) | 3 (10·3) |
| Five | 4 (10·8) | 1 (3·4) |
| Six | 1 (2·7) | 0 (0) |
| **Contexts** |  |  |
| Zero | 19 (51·4) | 11 (37·9) |
| One | 3 (8·1) | 4 (13·8) |
| Two | 4 (10·9) | 3 (10·3) |
| Three | 5 (13·5) | 7 (24·1) |
| Four | 3 (8·1) | 3 (10·3) |
| Five | 1 (2·7) | 0 (0) |
| Six | 0 (0) | 1 (3·4) |

Most definitions reference changes to pronouns (29/37) and names (29/37), while around half referenced changes to clothing (19/37) or hairstyle (16/37) (Table 2). Other practices included use of makeup for transfeminine individuals (2/37), verbally sharing their gender identity with others (i.e., “coming out”) (2/37) and the use of body modifiers, such as binders or packers (1/37). Five definitions did not specify any practices in their definition of social transition. The number of practices per definition varied widely (Table 4), with four practices being most common (14/37) followed by three practices (7/37).

Contexts in which social transition occurs (e.g. locations or with specific individuals such as friends) were mentioned in definitions across approximately half the studies (n=19) (Table 3), however the other half (n=18) did not specify any contexts in their definitions. The most commonly mentioned contexts were at home or with parents (14/37), or, at school (13/37). Less common contexts included in public (4/37), with friends (3/37), with extended family (2/37) and across all aspects of life (2/37). The number of contexts per definition of social transition varied considerably. Among the 19 studies that specified contexts, one (3/19), two (4/19), three (5/19), four (4/19) contexts were most commonly mentioned (Table 4).

### Measures of social transition

All identified measures of social transition are presented in Supplementary Table 2 (n=29 studies), while Tables 2, 3 and 4 summarise what is captured within these measures, focusing on the specific contexts and practices assessed. In six cases, an inclusion criterion was used as the measure.

Studies varied in their approach to measuring social transition. Only two of the 29 studies did not reference any specific practices; however, failure to reference specific contexts was relatively common (13/29). Some studies operationalised social transition as a change in a single practice (12/29) or context (4/29). Additionally, seven studies did not operationalise social transition with regard to specific practices or contexts, with six of these simply measuring whether participants “lived in role”. The measurement type also varied, with studies using binary measures (15/29), rating or Likert scales (7/29) and dimensional measures (7/29) that recorded data on multiple practices and/or contexts.

The most commonly measured practices were changes to pronouns (20/29), name (14/29), look or appearance (12/29), including specific mention of clothing (6/29) or hairstyle (7/29), and generally “living in role” (6/29). While some studies measured only one practice (12/29), others included multiple, such as two (4/29), three (7/29) or four (3/29).

Measured contexts included at school (13/29), at home or with parents (11/29), with friends (4/29), in “all contexts” (3/29) and online (3/29). Five additional contexts were measured in only one study each (e.g. with extended family, with strangers). Of the 29 studies, 11 did not specify any contexts, four focused on a single context, and the remainder (n=14) measured multiple contexts.

Four measurement tools for social transition were identified across the 29 studies: Gender Minority Stress and Resilience Measure (n=1),^57^ Social Transition Questionnaire (Trans20) (n=2),^58^ GIDS Service Questionnaire (n=1),^6^ and the Criteria for Full Transitions (n=1)^59^.

Evidently, no measurement tools were used consistently in the literature. Information on psychometric properties were unavailable, as most measures were derived from broader instruments rather than designed as standalone social transition scales. Where standalone scales were identified, no psychometric information were reported. None of these measures were co-designed with trans and gender diverse young people.

## Discussion

This scoping review revealed both the considerable variability in how social transition is defined and captured in the literature and the frequent oversimplification of this complex process. Social transition is inherently multifaceted, involving a range of practices that unfold across different social contexts, at different times, and at varying paces. However, despite this complexity, many studies reduce social transition to a single practice (e.g. pronoun or name change), while others fail to specify any practices at all. Similarly, the various contexts in which social transition can occur were inconsistently addressed. As a result, existing definitions and measures often provided a narrow or incomplete representation of an individual’s social transition, and the heterogeneity in existing approaches make comparison of findings about social transition across studies difficult.

The most commonly included practices were changes to pronouns (included in 78·4% of definitions, but 69·0% of measures), name (78·4% of definitions; 48·3% of measures), and clothing (51·4% of definitions; 20·7% of measures). Comparatively, the most commonly included contexts were at home/with parents (35·1% of definitions; 37·9% of measures), at school (32·4% of definitions; 44·8% of measures), and with friends (8·1% of definitions; 13·8% of measures), with these percentages noticeably lower than those for practices. This suggests that although the measures often tend to underrepresent a range of practices, contexts are commonly overlooked across both definitions and measures. These findings highlight an important gap currently missed in the literature: transitions that are occurring in different locations at different times (i.e. home vs school vs with friends). Therefore, even if mentioning many practices, researchers are unable to accurately measure where these practices are taking place.

Variations in the scope of these measures mean that studies may be capturing different constructs within social transition, potentially introducing significant bias into investigations of their associations with other variables, particularly mental health and wellbeing. This is concerning given ongoing debates regarding the safety and importance of early social transition for young people. For example, the Cass Review cites multiple studies included in this review^47,50,53^ to make the recommendation that social transition should be delayed or only partially undertaken in young people, as there is “no clear evidence that social transition in childhood has any positive or negative mental health outcomes”.^8^ (pp. 164) However, as highlighted by the authors of the systematic review commissioned for this section of the Cass Review, there are “extensive differences” between their included studies in how social transition was operationalised.^60^ These differences risk misrepresenting participant experiences, likely fail to capture the breadth of the real-life process of social transition, and may lead to inaccurate conclusions. Developing richer and more consistent measures would enable research to capture social transition and its association with other factors more accurately and comprehensively.

In addition to the heterogeneity observed, the measures identified in this review predominantly assume that a “full” or “complete” social transition is the goal for all individuals, as they do not account for an intention to only make specific changes, overlooking the diversity of experiences and transition goals among trans children and adolescents. This has likely resulted in the misclassification of some individuals, which is particularly concerning in studies using social transition status as the “intervention” under investigation. For example, young people who have met their transition goals with fewer gender-related changes than the measures expect may be classified as having a “partial transition”, despite no intention to undertake further changes (i.e. no intention to change their name).^61^ Conversely, some individuals who are classified as having undergone a “full transition” may still intend to make further changes, such as further alterations in appearance or transitioning in more social contexts. As the latter could foster a more stressful experience, studies using simplistic measures could misattribute this increased stress to undertaking more social transition steps. As mentioned prior, this misclassification is particularly problematic when comparing groups based on social transition status, as it may lead to misleading conclusions (i.e. false positive or false negative). To address these issues, future measures should account for an individual’s satisfaction with their current stage of social transition, as well as their intention for future changes in specific contexts.

Key aspects of social transition considered important by young people, such as having a safe and supportive environment for implementing social transition changes,^62^ were rarely present in the definitions and measures identified in this review. While these factors may not be central to defining social transition itself, they are crucial for understanding the circumstances in which these transitions occur. For example, current measures would likely conflate those who choose not to undertake specific aspects of social transition (e.g. name change) and those who feel unsafe to do so. The limited attention to contexts of social transition limits the understanding of how safe participants feel to make social transition changes. For example, online spaces (n=1 definition, 3 measures) can serve as safe environments for trans children and adolescents to explore their gender identity.^63^ Similarly, social transition with one’s extended family (n=2 definitions, 1 measure) may be an important context to measure, as this can be a context where individuals categorised as “fully” socially transitioned do not, or do not feel safe to, express their affirmed gender.^64^ These omissions highlight the need for individual context when measuring a young person’s social transition.

This scoping review provides a valuable summary of how social transition is defined and captured in the literature, particularly in the context of trans and gender diverse young people. Major strengths include the rigorous screening and extraction process, which was conducted by two reviewers at all stages to ensure the reliability of the data. Additionally, the inclusion of both qualitative and quantitative studies broadened the scope of the findings, resulting in richer data that offer a more nuanced understanding of the topic. The insights gained from this review lay an important foundation for future research into social transition, with the potential to refine both definitions and measures, ultimately enabling a more accurate account of the complex nature of social transition and its implications. However, there are also limitations to consider. First, the language used in this field is broad, evolving and often inconsistent across studies. As a result, some relevant studies may not have been captured, such as those referring to this process as social gender affirmation or gender affirmation more broadly, and those including social transition practices beyond those included in our search. Second, restricting the review to papers written in English likely limited the findings to a Western context, reflecting Western norms of gender identity and expression. Third, the review did not systematically assess how the identified measures were applied in studies, nor did it evaluate the evidence generated from their use, or potential subsequent biases in the findings, the latter of which falls outside the exploratory nature of a scoping review. Last, while the focused attention on children and adolescents in this review is a key strength, it also limits the generalisability of the findings to older populations or those who may undergo social transition at different stages of life.

## Conclusion and Recommendations

This review highlights the inconsistencies in how social transition has been defined and operationalised in existing research. Measures often fail to capture the dynamic and multi-faceted nature of social transition. To advance research in this area, future studies should:

1. **measure multiple practices** of social transition, including but not limited to, name, pronoun, and appearance change.
2. **measure the contexts** where young people have, and have not, undertaken each practice of social transition.
3. **measure young people’s desire to socially transition**, or to undertake more practices of social transition.
4. **collaborate with trans and gender diverse young people** in crafting a measure that accurately reflect the lived experiences of social transition.

These steps are likely to enhance the accuracy, validity, and applicability of future research to a real-world context.

## Author Contributions

WC led (for the updated search) data curation (literature search), project administration, writing (original draft), formal analysis/data analysis (in collaboration with the other first author), and visualisation (in collaboration with the other first author). WC also contributed to investigation, interpretation, and writing (review & editing). FL led (for original search) data curation (literature search), formal analysis, writing (original draft), formal analysis/data analysis (in collaboration with the other first author), and visualisation (in collaboration with the other first author). FL also contributed to conceptualisation, investigation, methodology, interpretation, supervision, project administration and writing (review and editing). HA contributed to conceptualisation, data curation (literature search – initial), and reviewed the final manuscript. MS contributed to data curation (literature search – both) and reviewed the final manuscript. CP contributed to conceptualisation and reviewed the final manuscript. PB contributed to writing (review & editing), interpretation, supervision and reviewed the final manuscript. SH contributed to data curation (literature search – updated) and reviewed the final manuscript. TD reviewed the final manuscript. KP contributed to conceptualisation, writing (review & editing), supervision and reviewed the final manuscript. MT contributed to conceptualisation, project administration, interpretation, writing (review & editing), supervision and reviewed the final manuscript. All authors had full access to the data in the study and had final responsibility for the decision to submit for publication. WC and FL were the authors responsible for accessing and verifying the underlying data.

## Data Sharing

The dataset generated for this study is available from the corresponding author upon reasonable request.

## Acknowledgements

We would like to thank Poh Chua from the Royal Children’s Hospital Library for her assistance in developing the search strategy, and Monsurul Hoq for his contribution in the initial stages of this project.

## Supplementary Material

**Ovid MEDLINE(R) Search <1946 to Sep 14, 2024>**

1 Transgender Persons/
2 Transsexualism/
3 “Sexual and Gender Minorities”/
4 Gender Dysphoria/
5 sex reassignment procedures/ or sex reassignment surgery/
6 Health Services for Transgender Persons/
7 exp gender identity/
8 (gender-identity or gender-dysphori* or gender-varian* or genderqueer* or queer* or intersex* or inter-sex* or transgender* or trans-gender* or transsexual* or trans-sexual or transexual* or two-spirit or trans-m#n or transm#n or trans-male* or transmale* or trans-wom#n or transwom#n or trans-female* or transfemale* or transfeminine or trans-feminine or transmasculine or trans-masculine or trans-folk* or transfolk* or trans-folx* or transfolx* or trans-people or transpeople or trans-person* or transperson* or trans-spectrum or AFAB or AMAB or cross-gender or QTPOC or M2F or MTF or F2M or FTM or gender-minorit* or sexual-minorit* or non-binary or nonbinary or gender-fluid* or genderfluid* or agender or gender-non-conform* or gender-nonconform* or TGNC or gender-ambigu* or gender-expression or gender-expansive or gender-diverse or transvesti* or third-gender or gender-creative or Gender-change or sex-reassignment or gender-reassignment or gender-confirmation or gender-affirmation or sex-change or sex-transformation or gender-transformation or gender-incongruen* or TGB or TGNB or TGDNB).tw,kf.
9 1 or 2 or 3 or 4 or 5 or 6 or 7 or 8
10 (boy or boys or girl or girls or child or children or childhood or pediatric* or paediatric* or school-age* or schoolage* or schoolchild* or schoolgirl* or schoolboy* or adolescen* or youth or youths or teen or teens or teenage*).af.
11 Social Participation/
12 (social-transition* or socially-transition* or ((pronoun? or look? or name?) and (social or famil* or school* or friend* or peer? or online))).tw,kf.
13 11 or 12
14 9 and 10 and 13
15 limit 14 to english language

**Supplementary Table 1:** Definitions and measures used to capture social transition in each included study, with these extracted as direct quotes from the original papers. Definitions are categorised by the scope of the definition (none, one context/type, or multiple types/contexts). Measures are categorised by study design (qualitative, quantitative longitudinal, or quantitative cross-sectional), informant (young person, parent, clinician), and data collection method (single question, rating scale, dimensional/multiple measures, or inclusion criterion). Definitions marked with an asterisk were derived from the study’s measurement approach rather than explicitly stated in text.

| Study | Scope of definition | Definition of social transition extracted from text | Study design, informant, and type of measure | Measure of social transition extracted from text |
| --- | --- | --- | --- | --- |
| **Provides both a definition and measure of social transition** |  |  |  |  |
| Bull, 2018<sup>19</sup> | Defined in more than one context or practice | *"Malpas (2011) defines social transition as a process that "consists of changing the child's social status and gender attributes—such as name, pronoun, clothing, official documentation, and registration in gendered activities and schools—in alignment with their gender identity, without making any definitive medical changes" (p. 456). In this study, we offer an amplification of that definition from the voices of the parents themselves. Participants spoke about two aspects of their child's social transition: small-t transition and capital-T transition (Participant 4). While participants described small-t transition is an ongoing process that has no clear beginning or end, they described capital-T transition as comprised of discrete moments of change such as when a child takes on a new name or the family begins to use different pronouns."* (p. 170) | Qualitative study | *"Some but necessarily all of the following means: (a) Change in name (e.g., named John at birth, transition to Joy); (b) Change in style of dress to facilitate expression of a different gender identity than was assigned at birth (e.g., wearing dresses and skirts instead of pants); (c) Change in hairstyle to express a different gender identity than was assigned at birth (e.g., letting hair grow long or braiding and using bows to facilitate a feminine expression, cutting hair short to facilitate a masculine expression); (d) Change in pronoun (going from he/him/his to she/her); (e) Changes must be expressed not just privately in the family home but rather made more public to the greater social community."* (p. 175) |
|  |  |  | Parent(s) |  |
|  |  |  | Inclusion criteria, binary measure. |  |
| De Castro, 2024<sup>20</sup> | Defined* in more than one context or practice | *Definition of social transition derived from the measure. | Quantitative, cross-sectional | *"A complete social transition was recorded when the transgender minor adopted the gender expression of their affirmed gender in their private, public and academic/professional lives... complete social transition (yes/no) and at what age"* (p. 36) |
|  |  |  | Medical Records |  |
|  |  |  | Binary measure |  |
| Durwood, 2021<sup>22</sup> | Defined in more than one context or practice. | *"Social transitions describe the process of changing one's name, pronouns, hairstyle, and clothing in order to allow the child to live as the gender with which a person identifies. Youth who have made childhood social transitions are necessarily highly supported in their gender identities at least by their parents, without the support of whom they would not have been able to make such a transition."* (p. 2) | Quantitative, cross-sectional | *"To be included in the present study, youth had to use the gendered pronouns stereotypically thought to be "opposite" their assigned sex at birth in all contexts, e.g., youth who were assigned male at birth and use "she/her" pronouns, or youth who were assigned female at birth and use "he/him" pronouns."* (p. 5) |
|  |  |  | Parent(s) |  |
|  |  |  | Inclusion criterion, binary question. |  |
| Durwood, 2022<sup>23</sup> | Defined in more than one context or practice | *"Changing hairstyles, clothing, name, and/or pronouns to live in line with their gender identities."* (p. 409) | Qualitative | *"To be included in the broader longitudinal study, youth needed to use the binary gendered pronouns other than the ones assigned to them at birth (e.g., a child assigned male at birth who used she/her pronouns, or a child assigned female at birth who used he/him pronouns; none used nonbinary pronouns at the start of the study) and be between the ages of 3–12 at the time that they first participated."* |
|  |  |  | Youth and/or parent |  |
|  |  |  | Inclusion criteria, binary measure |  |
|  |  |  |  | (p. 411) |
| Durwood, 2024<sup>24</sup> | Defined in more than one context or practice | “Social transitions describe the nonmedical process of changing one’s pronouns and often name, hairstyle, clothing, and so on to live as a different gender than the one assigned at birth.” (p. 984) | Quantitative, longitudinal | “We defined binary social transitions as youths having changed their pronouns to those stereotypically associated with the binary gender “opposite” their assigned sex at birth (e.g., for assigned male children, adopting “she/her” pronouns and for assigned female children, adopting “he/him” pronouns) across contexts. If youths changed their pronouns first in certain contexts (e.g., at home) and later in other contexts (e.g., at school), we considered the later of those times the age at which they socially transitioned.” (p. 986) |
|  |  |  | Parent(s) |  |
|  |  |  | Binary measure |  |
| Engel, 2023<sup>25</sup> | Defined in more than one context or practice | “...which includes coming out to others, using different names and pronouns, and changing one’s appearance.” (p. 2) | Quantitative, cross-sectional | “This tool was used to ask respondents to report the extent to which they had transitioned with respect to their name, pronouns, and appearance across different settings (home, school, and online). Young persons’ responses were categorised into fully transitioned, partially transitioned, and not transitioned”. (p. 4) |
|  |  |  | Young person |  |
|  |  |  | More than one procedure/context measured |  |
| Fast, 2018<sup>26</sup> | Defined in more than one context or practice | “When she was 5 years old, her parents allowed her to begin living as a girl in everyday life (meaning that they used the pronoun “she” and a new female name “Jazz,” but no medical or hormonal intervention occurred at that age)-a process called a social transition.” (p. 620) | Quantitative, cross-sectional | “Using the criteria for full transitions from Steensma, McGuire, Kreukels, Beekman, and Cohen-Kettenis (2013), participants had to be using the pronoun, clothing, and hairstyles associated with the “other” gender to count as socially transitioned.” (p. 625) |
|  |  |  | Young person |  |
|  |  |  | Inclusion criteria, binary measure |  |
| Gonzalez-Mendiondo, 2024<sup>27</sup> | Defined in more than one context or practice | Social transition is understood as the external recognition of the child’s lived gender identity, attendance to their needs, and exploring and supporting a process that entails ceasing to live socially according to the gender assigned at birth to live according to one’s gender identity, which may involve a change of name, pronouns, manner of presentation, hairstyle, or clothing”. (p. 1696) | Qualitative | “Who have made a complete binary social transition, accompanied by their families, to live according to their gender identity.” (p. 1697) |
|  |  |  | Parent(s) |  |
|  |  |  | Inclusion criteria, binary measure |  |
| Gulgoz, 2022<sup>28</sup> | Defined in a single context or practice | “Socially transitioned binary transgender children - i.e., they use “he” or “she” pronouns, but this gender does not align with their assigned sex - and a group of gender-non-conforming children - children who defy gender norms for their sex at birth, but either continue to use their original pronouns or have less binary identities (e.g., use “they” or similar non-binary pronouns).” (p. 3) | Quantitative, cross-sectional | “Which we had operationalized as using pronouns that are typically associated with the binary gender different from the one they were assumed to have at birth (i.e., an assigned female who uses “he” pronouns in all social contexts would be a trans boy in this study).” (p. 4) |
|  |  |  | Young person |  |
|  |  |  | Binary measure |  |
| Haywood, 2023<sup>6</sup> | Defined in more than one context or practice | “Social transition is often the first step a gender-diverse person takes to live as the gender that they identify with. Typically, this may involve choosing preferred pronouns, name, and/or aspects of appearance to reflect one’s experienced gender identity. This also | Quantitative, cross-sectional | “Social transition status is reported according to a combination of items. A full social transition in under 12s is defined as having changed their name informally or by deed poll, having asked others to use pronouns that reflect their gender identity, |
|  |  | *includes considering with whom, where, and in which situations they may feel comfortable to present as their identified gender.*" (p. 2) | Young person and parent(s)<br>More than one procedure/context measured | *and having their caregivers define their gender identity as fixed and different to their gender assigned at birth (this included identifying as non-binary). As those 12 + years completed their own questionnaires, additional items included: having permanently changed their appearance in accordance with their current gender identification with family, friends, at school, with strangers, and online. Parental reports querying gender fluidity are also not asked in the 12 + questionnaires. Young people that report some but not all of these aspects of social transition are categorised as partially socially transitioned.*" (p. 3) |
| Kuper, 2019<sup>30</sup> | Defined* in more than one context or practice | *Definition of social transition derived from the measure. | Quantitative, cross-sectional<br>Young person (from clinician notes)<br>More than one procedure/context measured | *Clinicians reported information obtained from participants on: "(a) whether they had initiated changes in clothing, appearance (mainly hairstyle but also use of makeup and/or jewellery for affirmed females), and name/pronoun (yes/no), and if so, at what age, (b) whether they had disclosed their gender identity to parents, extended family, friends, and teachers/staff (yes/no), and if so, at what age and whether the reaction was described as distressing in any way (yes/no)." (p. 220)* |
| Kuvalanka, 2017<sup>32</sup> | Defined in more than one context or practice | *"Changing their first names, pronouns, hair, and clothing to better align with their gender identities."* (p. 890) | Quantitative, cross-sectional<br>Parent(s)<br>Binary measure | *"They had switched pronouns from "he" to "she" or vice versa, and they (or their parents) asked everyone in their lives to make the switch"* (p. 894) |
| Lane, 2025<sup>34</sup> | Defined* in more than one context or practice | *Definition of social transition derived from the measure. | Quantitative, cross-sectional<br>Medical records<br>Rating (Likert scale) measure | *Social transition was coded as no (no indication of social transition), partial (some indication of social transition but in limited contexts or only applied to certain gender expressions), or full (name, pronoun, and appearance change to a gender other than that registered at birth in all contexts). (p. 935)* |
| Levitan, 2019<sup>35</sup> | Defined in more than one context or practice | *"...the degree of variation in gender presentation among adolescents, in how far they have 'outed' themselves to others, or live in their preferred gender role."* (p. 1488) | Quantitative, cross-sectional<br>Young person<br>Rating (Likert scale) measure | *"Social transitioning was measured using a self-constructed item that assessed the degree to which adolescents had undergone a complete social transition or lived in their preferred gender role in three different life areas, respectively: at home, with friends/in their peer group, and at school. Since multiple answers were possible, the combination of the different answers were recoded as follows: 1 = "no social transition, yet (living in birth-assigned gender role)"; 2 = "partial social transition in at least 1 out of 3 life areas (or in-between gender roles)"; 3 = almost complete social transition (living in the new gender role in at least 2 out of 3 life areas; e.g., at home and with friends, but not in school)"; 4 = "complete social transition in all life areas (living in the new gender role of the preferred gender at home, with friends/peers and at school)". Resulting in a four-point scale, this measure allows for the assessment of the gradual degree to which a child/adolescent has transitioned to another gender role, socially."* (p. 1490-1) |
| Morandini, 2022<sup>36</sup> | Defined in more than one context or practice | *"Had changed their clothing, hairstyle, name, and pronouns to match their gender identity."* (p. 481) | Quantitative, cross-sectional | *"Living in role, which referred to whether young people were currently living in their preferred gender role (e.g. adopting gender markers including clothes and hair styles of their felt gender) was divided into fulltime, part-time and not living in role. Name change, which referred to whether young people had adopted a name and pronouns congruent with their felt gender, was divided into yes (full-time), partly (part-time) and no (had not adopted a new name)."* (p. 485) |
|  |  |  | Young person |  |
|  |  |  | Rating (Likert scale) measure |  |
| Morandini, 2023<sup>37</sup> | Defined in more than one context or practice | *"i.e., living in their affirmed gender rather than their gender assigned at birth, which may involve changing their physical gender markers such as hair and clothing as well as their name and gender pronouns."* (p. 1045) | Quantitative, cross-sectional | *"The key variables of social transition, "living in birth- assigned gender versus affirmed gender role" and "name change," were originally rated in three categories: "no" (i.e., no social transition/name change), "partly" (i.e., partial social transition/name change), or "yes" (i.e., full social transitioned/name change)" (consistent with how social transition was assessed by Steensma et al., [2013]). Because very few respondents were rated as partially socially transitioned or as partially using a new name, this was not a viable cell for analysis. We, therefore, combined partially and fully transitioned patients into a single group, creating dichotomous variables for both living in role and name change (0 = No transition or No name change, 1 = Any social transition or Any name change)."* (p. 1049-50) |
|  |  |  | Young person |  |
|  |  |  | Rating (Likert scale) measure, analysed binary |  |
| O'Bryan, 2018<sup>38</sup> | Defined* in more than one context or practice | *Definition of social transition derived from the measure. | Quantitative, cross-sectional | *"Gender identity presentation (i.e., pronoun use, use of preferred name, dress) both at home and in public (i.e., school or workplace)."* (p. 183) |
|  |  |  | Young person |  |
|  |  |  | Binary measure |  |
| Olson, 2016<sup>39</sup> | Defined in more than one context or practice | *"Involve changes in the child's appearance (e.g. hair, clothing), the pronoun used to refer to the child, and typically also a change in the child's name."* (p. 3) | Quantitative, cross-sectional | *Social transition comprised the following criteria: "(1) identify as the gender "opposite" their natal sex in everyday life (i.e. they identified as male or female, but not the gender that aligned with their sex at birth), (2) present in all contexts (e.g. at school, in public) as that gender identity, (3) use the pronoun matching their gender rather than their natal sex..."* (p. 3) |
|  |  |  | Young person |  |
|  |  |  | Inclusion criteria, binary measure |  |
| Olson, 2018<sup>40</sup> | Defined in more than one context or practice | *"Changing their pronouns and first names to match their gender identity while still in the prepubescent years"* (p. 1) | Quantitative, cross-sectional | *"Social transitions were defined as occurring if the child used the pronoun of the gender "opposite" their natal sex in all contexts (e.g., at home, at school, upon introduction to strangers)."* (p. 4) |
|  |  |  | Young person |  |
|  |  |  | Binary measure |  |
| Rae, 2019<sup>43</sup> | Defined* in a single context or practice | *Definition of social transition derived from the measure. | Quantitative, longitudinal | *Children were defined as having socially transitioned if they "changed pronouns to align with the gender opposite their assigned sex (e.g., an assigned male going by "she"; Fast & Olson, 2018; what is called a "complete transition" by Steensma et al., 2013, p. 585). By the time children change their pronouns, or at the same time, they typically change their first name (if their original first name was gendered), hairstyle, and clothing. To assess later transition status, we contacted parents for* |
|  |  |  | Young person |  |
|  |  |  | Binary measure | confirmation, or we received an update via an in-person follow-up visit or parental online survey. If there had been multiple contacts after the initial data collection, the most recent contact before the article submission was used to determine whether the child had socially transitioned.”<br>(p. 671) |
| Renley, 2022<sup>44</sup> | Defined in more than one context or practice. | “Changing name and pronouns can be a part of social transitioning, the process where an individual begins to socially present as the gender that aligns with one’s gender identity (Rafferty et al., 2018).”<br>(p. 781) | Quantitative, cross-sectional<br>Young person | “To measure the frequency that adults and students use their correct pronouns, youth were asked “At school, do adults and students call you by the pronouns (e.g., she, her, hers) that you want to be called?” Participants selected from the response options of Never, Occasionally, Sometimes, Most of the time, and Always. The item was scored on a scale of 0 (Never) to 4 (Always), with higher scores indicating greater frequency of correct pronoun use.<br>To measure whether adults and students refer to youth by their correct names, youth responded to a single item that asked “At school, do adults and students call you by the name that you want to be called?” with response options of Never, Occasionally, Sometimes, Most of the time, and Always. The item was scored on a scale of 0 (Never) to 4 (Always), with higher scores indicating greater frequency of correct name use.<br>To determine whether youth were able to use the bathrooms/ locker rooms that match their gender at school, youth were asked “At school, do you use restrooms and locker rooms that match your gender identity?” with the following response options: Never, Occasionally, Sometimes, Most of the time, and Always. The item was scored on a scale of 0 (Never) to 4 (Always), with higher scores indicating greater use of bathrooms/locker rooms that match the respondent’s gender.”<br>(p. 784) |
|  |  |  | Rating (Likert scale) measures |  |
| Sievert, 2021<sup>47</sup> | Defined in more than one context or practice | “Living in the preferred gender role in the following three everyday life areas: at home/with family, with friends/peers, and at school.”<br>(p. 80) | Quantitative, cross-sectional | “The degree to which a child had transitioned socially was assessed via three items, asking whether they lived in their preferred gender role in the following three everyday life areas:<br>(1) at home/with family, (2) with friends/peers, and (3) at school. Taking into consideration that multiple answers were possible, the final variable was summed up and then recoded as 1 (no social transition, yet, and living in birth-assigned gender role); 2 (partial social transition in at least 1 out of 3 life areas or in between gender roles); 3 (almost complete social transition, living in the new gender role in at least 2 out of 3 life areas); 4 (complete social transition in all life areas). In addition, parents were asked on a scale from 1 (not at all) to 5 (completely) in how far they supported their child’s current gender role or social transition.”<br>(p. 84) |
|  |  |  | Parent(s) |  |
|  |  |  | Rating (Likert scale) measure |  |
| Steensma, 2013<sup>50</sup> | Defined* in more than one | *Definition of social transition derived from the measure. | Quantitative, longitudinal | “Social role transition was determined through two questions by one of the parents around the time of referral. Parents indicated whether their child had socially transitioned to the preferred gender role on a 3-point scale; no; yes, but not in all |
|  | context or practice |  | Young person | situations; or yes, completely. In an open question, they could give further information on hairstyle, clothing, and in which pronoun and name the child was addressed. Based on this information, the children were categorized as follows: no social transition; partial transition (transition in clothing style and hairstyle, but without a change of name and pronoun change); complete transition (transition in clothing and hairstyle; change of name and use of pronoun). Because of the unequal distribution over the 3 categories for boys and girls, the 3-point scale was recoded into a dichotomous scale in which 0 indicated no transitioning (category 1) and 1 indicated some transitioning (categories 2 and 3). (p. 584-5) |
|  |  |  | Rating (Likert scale) measure |  |
| Thoma, 2023<sup>51</sup> | Defined in more than one context or practice | "Including disclosing gender identity to parents and peers, using pronouns and a name that align with their true gender identity, and changing their hairstyle or clothing" (p. 2) | Quantitative, cross-sectional | Engagement in gender transition steps: "Dressing in a different way", "Doing hair in a different way" and "Going by a different name". (p. 16) |
|  |  |  | Young person |  |
|  |  |  | More than one procedure/context measured |  |
| Tollit, 2024<sup>52</sup> | Defined in more than one context or practice | "...(*i.e.*, changing the way they present themselves), and this can vary in terms of methods (*e.g.*, through dress, name, pronouns) and environments in which this occurs (*e.g.*, home, school, online).” (p. 2) | Quantitative, cross-sectional | “The Social Transition Questionnaire measures the extent to which a person has socially transitioned in different contexts (*i.e.*, name, pronouns, appearance) and settings (*i.e.*, home, school, online).” (p. 4) |
|  |  |  | Young person and parent(s) |  |
|  |  |  | More than one procedure/context measured |  |
| Wittlin, 2025<sup>54</sup> | Defined in more than one context or practice | “...*changed their names, pronouns, clothing, and/or hairstyles to align with their gender identity.*” (p. 228) | Quantitative, longitudinal | "were assigned male at birth, living as girl, and using she/her pronouns or assigned female at birth, living as a boy, and using he/him pronouns; further details about recruitment are provided in Gulgoz et al. (2019)” (p. 230)<br>Reference: "This means that, across every context (at home, at school, meeting new people), they were using a binary pronoun (*i.e.*, he or she) that was not associated with their sex assigned at birth."<sup>56</sup> (Supplementary Material, p. 8) |
|  |  |  | Young person and parent(s) |  |
|  |  |  | More than one procedure/context measured |  |
| Provides only a definition of social transition |  |  |  |  |
| Aramburu-Algeria, 2018<sup>17</sup> | Defined in more than one | “Living in their affirmed gender and using their chosen name and pronouns at home, at school, with extended family, and in society.” | Qualitative study | Social transition explored in qualitative interviews, rather than having a strict measure. |
|  |  |  | Parent(s) |  |
|  | context or practice | (p. 133) | Inclusion criterion, not operationalised |  |
| Bocek, 2024<sup>18</sup> | Defined in more than one context or practice | *"Nonmedical gender-affirming interventions, which are often referred to as social transition, may include name change, voice coaching, wearing affirming clothing, or coming out at home or school."*<br>(p. 1) | Quantitative, cross-sectional<br>Young person<br>Measured related concepts | Measured related concept, desired social transition, rather than practices and contexts where changes had been enacted by the young person. |
| Durwood, 2017<sup>21</sup> | Defined in more than one context or practice | *"A nonmedical decision to allow a child to change his or her first name, pronouns, hairstyle, and clothing to live everyday life as one's asserted gender."*<br>(p. 2) | Quantitative, cross-sectional<br>Young person<br>Inclusion criteria, not operationalised |  |
| Horton, 2023<sup>29</sup> | Defined in more than one context or practice | *"Pronoun change in a majority of settings (i.e., home and school) was taken as the key indicator of what is known as a social transition."*<br>(p. 75) | Qualitative study<br>Parent(s)<br>Explored qualitatively, no measure | Social transition explored in qualitative interviews, rather than having a strict measure. |
| Kuvalanka, 2014<sup>31</sup> | Defined in more than one context or practice | *"Present their true gender selves to the outside world. Time period during which the child outwardly changes gender through, for example, name, hairstyle, and clothing changes."*<br>(p. 357) | Qualitative<br>Parent(s)<br>Explored qualitatively, no measure | Social transition explored in qualitative interviews, rather than having a strict measure. |
| Olson, 2019<sup>41</sup> | Defined in more than one context or practice | Social transition occurred *"if a child was using binary pronouns (he or she) consistently in all environments (e.g., at school, home, with strangers) and that pronoun contrasted with the child's pronoun at birth"*.<br>(p. 231) | Qualitative<br>Parent(s)<br>Explored qualitatively, no measure | Social transition explored in qualitative interviews, rather than having a strict measure. |
| Olson, 2022<sup>42</sup> | Defined in a single context or practice | *"A 'complete' binary social transition included changing their pronouns to the binary gender pronouns that differed from those used at their births."*<br>(p. 2) | Quantitative, longitudinal<br>Young person<br>Inclusion criteria, no measure |  |
| Schimmel-Bristow, 2018<sup>45</sup> | Defined in more than one context or practice | *"Refers to coming out as transgender; using affirmed name and pronouns; expressing gender identity through clothing, make-up, body modifiers (e.g., binders, packers, bra stuffing), and other similar forms of expression"*<br>(p. 274) | Qualitative<br>Youth and parent(s)<br>Explored qualitatively, no measure | Social transition explored in qualitative interviews, rather than having a strict measure. |
| Shim, 2020<sup>46</sup> |  | *Self-disclosure of publicly identifying along the transmasculine spectrum (for assigned females).* | Quantitative, cross-sectional |  |
|  | Defined in a single context or practice | (p. 525) | Young person<br>No measure |  |
| Wong, 2019<sup>55</sup> | Defined in more than one context or practice | *“Social gender transition (SGT) entails changes to characteristics such as the child’s name, pronouns, and appearance (e.g., hairstyle, clothing) in all aspects of day-to-day life, whereas a partial SGT consists of either some of these changes and/or these changes only taking place in certain contexts (e.g., in the child’s home, but not in public places such as in the child’s school.”*<br>(p. 242) | Quantitative, cross-sectional<br>Parent(s)<br>No measure |  |
| van der Vaart, 2022<sup>53</sup> | Defined in more than one context or practice | *Considered to be equivalent to the process of a child living in their experienced gender. Children who presented themselves using a name and pronouns and, or, wearing clothing and have hairstyle incongruent to their assigned gender at birth were categorized as children living in their experienced gender.*<br>(p. 1080) | Quantitative, cross-sectional<br>Young person and parent(s)<br>Explored qualitatively, no measure | Social transition explored in qualitative interviews, rather than having a strict measure. |
| **Provides only a measure of social transition** |  |  |  |  |
| Legrange, 2024<sup>33</sup> |  |  | Quantitative, cross-sectional<br>Medical records<br>More than one procedure/context measured | *“We also investigated aspects of social transition, such as the affirmation of gender at school, within the family, and among peers, as well as any changes in legal identification (name and gender marker). Notably, according to French law, a change of name is possible at local town halls without a minimum age requirement, with the agreement of both parents. Gender changes on official documents are possible from the age of 18 years (or 16 years if the teenager is legally emancipated).”*<br>(p. 4) |
| Sood, 2021<sup>48</sup> |  |  | Quantitative, cross-sectional<br>Young person<br>Binary measure | *Social transition status was captured by a single yes/no question from the Gender Minority Stress and Resilience Measure: “Do you currently live in your affirmed gender all or almost all of the time? Your affirmed gender is the one you see as accurate for yourself.”*<br>(p. 1137) |
| Sorbara, 2020<sup>49</sup> |  |  | Quantitative, cross-sectional<br>Young person and parent(s)<br>Binary measure | *“The age at which the youth first social transition in at least one real-life environment.”*<br>(p. 3) |

